# Vascular risk is not associated with PET measures of Alzheimer’s disease neuropathology among cognitively normal older adults

**DOI:** 10.1101/2021.09.01.21256275

**Authors:** Murat Bilgel, Alisa Bannerjee, Andrea Shafer, Yang An, Susan M. Resnick

## Abstract

Cardiovascular disease (CVD) is associated with a higher risk of developing dementia. Studies have found that vascular risk factors are associated with greater amyloid-β (Aβ) and tau burden, which are hallmark neuropathologies of Alzheimer’s disease (AD). Evidence for these associations during the preclinical stages of AD, when Aβ and tau pathologies first become detectable, is mixed. Quantifying the effect of vascular risk among cognitively normal individuals can help focus the efforts to develop therapeutic approaches aimed at modifying the course of preclinical AD.

Using Bayesian analysis, we examined the relationship of Aβ and tau pathology with concurrent vascular risk among 87 cognitively normal individuals (median age 77, interquartile range 70– 83) in the Baltimore Longitudinal Study of Aging. We quantified vascular risk as the probability of developing CVD within 10 years using published equations from the Framingham Heart Study. Aβ and tau pathologies were measured using positron emission tomography.

As expected, Aβ positive participants had greater tau in the entorhinal cortex (EC) and inferior temporal gyrus (ITG) (difference in means = 0.09, p < 0.05 for each region), and 10-year CVD risk was positively correlated with white matter lesion burden (r = 0.24, p = 0.03). However, we did not find any associations between CVD risk and Aβ or tau. The data provided over two- and four-fold evidence towards the lack of a correlation between CVD risk and tau in the EC (Bayes factor BF = 2.4) and ITG (BF = 4.0), respectively. We found over three-fold evidence towards the lack of a difference in mean CVD risk by Aβ group (BF = 3.4). These null findings were replicated using a data-driven vascular risk score in the BLSA based on a principal component analysis of eight indicators of vascular health.

Our data provide moderate evidence towards the lack of an association between vascular risk and concurrent AD neuropathology among cognitively normal older adults. This finding suggests that vascular risk and AD neuropathology may constitute independent pathways in the development of cognitive impairment and dementia.

**Highlights:**

- Vascular risk is associated with white matter lesion load, but not amyloid-β or tau
- Moderate evidence for lack of association between vascular risk & early AD pathology
- Vascular risk and Alzheimer’s neuropathology may constitute independent pathways

## 1 Introduction

Alzheimer’s disease (AD) and cardiovascular disease (CVD) are both growing public health issues. About 5.8 million Americans over the age of 65 are currently living with AD. As the overall US population ages, these numbers are expected to increase dramatically (Alzheimer’s Association, 2020). Several studies have shown that cerebrovascular disease, including history of stroke and infarcts, may increase the risk of developing clinical symptoms of AD and dementia among older adults (Gamaldo et al., 2006; Schneider et al., 2004; Troncoso et al., 2008), and often occurs concurrently with AD neuropathology, namely, amyloid-β (Aβ) plaques and neurofibrillary tau tangles (Azarpazhooh et al., 2018; Gorelick et al., 2011; Kapasi et al., 2017). Vascular risk factors, such as hypertension, hyperlipidemia, hypertriglyceridemia, diabetes, obesity, and stroke, are thought to promote vascular damage, which directly promotes Aβ accumulation and directly and indirectly promotes tau accumulation (Zlokovic, 2011). Therefore, understanding how vascular risk may contribute to the development and progression of AD neuropathology has become increasingly important.

While the relationship between vascular risk and dementia has been widely acknowledged (Livingston et al., 2020), evidence for the contribution of vascular risk to the early neuropathological manifestation of AD while individuals are cognitively normal remains mixed. Mid-life, rather than late-life, vascular risk appears to be more closely associated with Aβ deposition among individuals without dementia (Gottesman et al., 2017), and this association might be dependent on the presence of at least one *APOE* ε4 allele (Jeon et al., 2019; Rodrigue et al., 2013). Among cognitively normal individuals, greater tau burden has been associated with higher triglycerides in midlife (Nägga et al., 2018), insulin resistance (Laws et al., 2017), obesity and hypertension (Bos et al., 2019), and overall CVD risk (Bos et al., 2019; Rabin et al., 2019). Interestingly, a longitudinal study of cognitively normal individuals found that reducing mean arterial pressure (MAP) was associated with an increase in CSF p-tau_181_ in the hypertensive group (Glodzik et al., 2014). Both Aβ (Rabin et al., 2019) and medication use for vascular risk (Köbe et al., 2020) have been suggested as moderators of the relationship between CVD risk and tau. In contrast, several studies of cognitively normal individuals reported a lack of an association between vascular risk and Aβ (Bos et al., 2019; Glodzik et al., 2014; Laws et al., 2017; Marchant et al., 2012; Pettigrew et al., 2020; Rabin et al., 2018; Vassilaki et al., 2016; Vemuri et al., 2015; Walters et al., 2018) or tau (Köbe et al., 2020; Pettigrew et al., 2020), but none of these studies quantified the degree of evidence towards the absence of an association.

Determining the extent of the association between vascular risk and early AD neuropathology can help inform strategies to prevent or delay future cognitive decline. If vascular risk is a player in the development of AD neuropathology, early intervention on modifiable vascular risk factors could be critical in reducing the risk of mild cognitive impairment (Williamson et al., 2019) or dementia. On the other hand, if vascular risk factors are not associated with early AD neuropathology, this finding would suggest that vascular risk and AD neuropathology are separate pathways in preclinical AD. In either case, this knowledge would help better focus the efforts to develop therapeutic approaches aimed at modifying the course of preclinical AD.

In this study, we examined the relationship of Aβ and tau pathology with concurrent vascular risk among 87 cognitively normal individuals in the Baltimore Longitudinal Study of Aging (BLSA). We quantified vascular risk using two approaches: first, we calculated the probability of developing CVD within 10 years using previously published equations from the Framingham Heart Study, and second, we constructed a data-driven vascular risk score in the BLSA based on eight variables that are established indicators of vascular risk. Aβ and tau pathologies were measured using positron emission tomography (PET). Based on previous literature, we hypothesized that individuals with higher vascular risk would have greater levels of Aβ and tau pathology.

## 2 Materials and Methods

### 2.1 Participants

We used data for cognitively normal participants in the BLSA who had an ^18^F-flortaucipir (FTP) PET scan to measure phosphorylated tau burden, ^11^C-Pittsburgh compound B (PiB) PET scan to measure fibrillar Aβ burden, magnetic resonance imaging (MRI) scans to quantify total white matter volume (WMV) and total white matter lesion load (WML), and measurements for vascular risk variables (described below). We used measurements closest in time to the first FTP PET scan. All measurements were within 3.3 years of the baseline FTP PET. We excluded participants who had missing values for any of these variables. Additionally, one participant with outlying FTP SUVR values was excluded.

Cognitively normal status was based on either (1) a Clinical Dementia Rating (Morris, 1993) of zero and ≤3 errors on the Blessed Information-Memory-Concentration Test (Fuld, 1978), and therefore the participant did not meet criteria for consensus conference; or (2) the participant was determined to be cognitively normal based on thorough review of clinical and neuropsychological data at consensus conference. At enrollment into the PET neuroimaging substudy of the BLSA, all participants were free of central nervous system disease (dementia, stroke, bipolar illness, epilepsy), severe cardiac disease, severe pulmonary disease, and metastatic cancer.

Research protocols were conducted in accordance with United States federal policy for the protection of human research subjects contained in Title 45 Part 46 of the Code of Federal Regulations (45 CFR 46), approved by local institutional review boards (IRB), and all participants gave written informed consent at each visit. The BLSA PET substudy is governed by the IRB of the Johns Hopkins Medical Institutions (protocol numbers NA_00051793 and IRB00047185), and the BLSA study is overseen by the National Institute of Environmental Health Sciences IRB.

### 2.2 Vascular risk variables

All participants enrolled in the BLSA-NI study received measurements of high density lipoprotein (HDL) cholesterol, low-density lipoprotein (LDL) cholesterol, total cholesterol, triglyceride level, body-mass index (BMI), and systolic and diastolic blood pressure (SBP and DBP) at the time of their BLSA visit. Mean arterial pressure (MAP) was computed using SBP and DBP.

The presence of hypertension, diabetes, and high cholesterol, as diagnosed by a doctor or other health professional, was determined via self-report through a medical history interview conducted during the participant’s BLSA visit. Self-reported medication use for hypertension, diabetes, and high cholesterol was also determined through this questionnaire. For each of these conditions, participants were considered to have the condition if they reported medication use for the condition; otherwise, their self-reported diagnosis was used. If self-reported diagnosis was not available, we considered participants to have the condition if they had suprathreshold measurements (SBP ≥ 130 mmHg or DBP ≥ 80 mmHg for hypertension, fasting glucose > 125 mg/dL for diabetes, total cholesterol ≥ 200 mg/dL for high cholesterol), and not to have the condition otherwise. Smoking status was determined through a self-reported questionnaire and categorized as current smoker, former smoker, or never previously smoked.

We calculated a 10-year CVD risk score using the equations reported by D’Agostino et al. based on the Framingham Heart Study (D’Agostino et al., 2008). This risk score incorporates age, sex, SBP, treatment for high blood pressure, total cholesterol, diabetes diagnosis, and current smoking, and reflects the probability of developing CVD (i.e., coronary heart disease, cerebrovascular disease, peripheral vascular disease, and heart failure) within 10 years. Missingness in self-reported anti-hypertensive drug use was considered as no anti-hypertensive drug use.

### 2.3 MR imaging

MPRAGE (repetition time [TR] = 6.8 ms, echo time [TE] = 3.2 ms, flip angle = 8°, image matrix = 256 × 256 × 170, voxel size = 1 × 1 × 1.2 mm^3^) and FLAIR (TR = 11 s, TE = 68 ms, inversion time = 2800 ms, image matrix = 240 × 240 × 150, voxel size = 0.83 × 0.83 × 3 mm^3^) scans were acquired on a 3 T Philips Achieva scanner. MPRAGE scans were used to compute anatomical labels and regional brain volumes with Multi-atlas region Segmentation using Ensembles of registration algorithms and parameters (Doshi et al., 2016). FLAIR scans were used to segment white matter lesions with DeepMRSeg (Doshi et al., 2019).

### 2.4 PET imaging

To assess phosphorylated tau burden, FTP PET scans were acquired over 30 min on a Siemens High Resolution Research Tomograph (HRRT) scanner starting 75 min after an intravenous bolus injection of approximately 370 MBq of radiotracer. Dynamic images were reconstructed using ordered subset expectation-maximization to yield 6 time frames of 5 min each with approximately 2.5 mm full-width at half-maximum (FWHM) at the center of the field of view (image matrix = 256 × 256 × 107, voxel size = 1.22 × 1.22 × 1.22 mm^3^). Following time frame alignment, the 80–100 min average PET image was partial volume corrected using the region-based voxelwise method (Thomas et al., 2016). The corrected image was then used to compute standardized uptake value ratio (SUVR) images with inferior cerebellar gray matter as the reference region. FTP PET image analysis workflow is described in further detail in Ziontz et al. (Ziontz et al., 2019). We computed the average bilateral SUVR in the entorhinal cortex (EC) and the inferior temporal gyrus (ITG).

To assess fibrillar Aβ burden, PiB PET scans were obtained over 70 min on either a GE Advance or a Siemens HRRT scanner immediately following an intravenous bolus injection of approximately 555 MBq of radiotracer. Scans acquired on the GE Advance were reconstructed using filtered backprojection with a ramp filter to yield 33 time frames with approximately 4.5 mm FWHM at the center of the field of view (image matrix = 128 × 128 × 35, voxel size = 2 mm × 2 mm × 4.25 mm). Scans acquired on the Siemens HRRT were reconstructed using ordered subset expectation-maximization to yield 33 time frames with approximately 2.5 mm FWHM at the center of the field of view (image matrix = 256 × 256 × 207, voxel size = 1.22 mm × 1.22 mm × 1.22 mm). HRRT scans were smoothed with a 3 mm FWHM isotropic Gaussian kernel and then resampled to bring their spatial resolution closer to that of the GE Advance scans and match their voxel size. Following time frame alignment and co-registration with MRI, distribution volume ratio (DVR) images were computed using a spatially constrained simplified reference tissue model with cerebellar gray matter as the reference region (Zhou et al., 2007). Mean cortical Aβ burden was calculated as the average of the DVR values in cingulate, frontal, parietal (including precuneus), lateral temporal, and lateral occipital cortical regions, excluding the sensorimotor strip. PiB PET image analysis workflow is described in further detail in Bilgel et al. (Bilgel et al., 2019). Leveraging longitudinal PiB PET data available on both GE Advance and HRRT scanners for 79 BLSA participants, we estimated the parameters of a linear model mapping mean cortical DVR values between the GE Advance and HRRT scanners and applied this mapping to all HRRT values to harmonize them with the GE Advance values. Individuals were categorized as PiB –/+ based on a mean cortical DVR threshold of 1.06, which was derived from a Gaussian mixture model fitted to harmonized mean cortical DVR values at baseline.

### 2.5 Statistical analyses

#### 2.5.1 Sanity checks

We first assessed if CVD risk was associated with log-transformed total WML to total WMV ratio using Pearson’s correlation coefficient. We calculated the 95% confidence interval (CI) for the correlation coefficient based on Fisher’s *z*-transform. This analysis was intended as a validation of the CVD risk variable in our sample. We expected to find a positive correlation.

Next, we assessed if FTP SUVRs in the EC and the ITG were associated with Aβ status using two-sample t-tests allowing for unequal variance. This analysis was intended as a validation of regional FTP SUVR measures. We expected to find higher FTP SUVR among PiB+ participants. We also conducted a correlation analysis using continuous mean cortical PiB DVR instead of dichotomous Aβ status.

We did not adjust for age or sex in these sanity check analyses because they are factored into the CVD risk calculation and we did not want to remove any age- or sex-related association.

#### 2.5.2 Main analysis

Our main analysis involved assessing the Pearson’s correlation between CVD risk score and regional FTP SUVRs. To assess the degree of evidence provided by our data for the null hypothesis that there is no correlation between CVD risk score and FTP SUVR, we computed Bayes factors (BFs) as the ratio of the evidence for the null to the evidence for the alternative hypothesis. For the correlation parameter, we used a prior with a stretched Beta (1/*k*, 1/*k*) distribution (Ly et al., 2016). We set the scale parameter *κ* to 1/3 for a medium-width prior. We investigated the impact of the scale on BFs by varying *κ* between 0.001 (which yields a prior highly concentrated around 0, favoring low correlation values) and 1 (which yields a uniform prior).

We evaluated whether mean CVD risk differs between Aβ groups using a two-sample t-test allowing for unequal variance. To assess the degree of evidence provided by our data for the null hypothesis that there is no difference in means, we computed BFs using a Jeffreys prior on the variance^*^ and a Cauchy prior on the standardized effect size (Ly et al., 2016). The scale of the Cauchy prior was set to 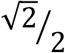 for the main analysis for a medium-width prior. We investigated the impact of the scale on BFs by varying it between 0.001 (which yields a prior highly concentrated around 0, favoring very small differences in group means) and 1.5 (which yields a wide Cauchy distribution). Additionally, we performed a correlation analysis using continuous mean cortical PiB DVR instead of dichotomous PiB group.

Finally, we calculated the BFs corresponding to the comparison of a linear regression model for FTP SUVR that included CVD risk, *APOE* ε4 status, and Aβ group as independent variables versus a linear regression model that included only *APOE* ε4 status and Aβ group as independent variables, with the latter model serving as the null hypothesis that the regression coefficient for CVD risk is equal to 0. We also investigated the inclusion of an Aβ group by CVD risk interaction term in the linear regression model. We did not consider age or sex as covariates in these regression models since they are already factored into the CVD risk calculation. The joint prior for the intercept and error variance was assumed to be proportional to the inverse of the error variance, and the regression coefficients were assumed to have a Zellner’s *g*-prior, with *g* having an Inv-Gamma(1/2, *r*/2) distribution (Liang et al., 2008). The scale parameter *r* was set to 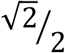 for a medium-width prior.

We interpreted BFs (computed with the null on the numerator) between 1 and 3 as weak evidence, and between 3 and 10 as moderate evidence towards the null hypothesis that there is no association between CVD risk and the variable investigated, following the guidance in Quintana & Williams (Quintana and Williams, 2018).

#### 2.5.3 Analysis using principal component scores

To eliminate the possibility that 10-year CVD risk may not be the ideal summary measure of vascular risk in our sample, we constructed summary scores based on BLSA data and repeated our sanity checks and main analysis using these scores instead of CVD risk. To define a study-specific vascular risk variable, we performed principal component analysis (PCA) on BMI, MAP, HDL, LDL, total cholesterol, triglycerides, fasting glucose, smoking status (current smoker vs. not). First, we created a cross-sectional sample of all BLSA participants who had measurements for all of these 8 variables. To verify the replicability of the principal components (PCs), we split this sample into training and testing sets, and performed PCA separately in each data set. Details are described in the Supplementary Material. The first three PCs computed in the training set were used to calculate scores for each of the 87 participants included in this study. Scores along each of the PCs were then used in place of CVD risk to repeat the correlation tests with log-transformed total WML ratio, as described in 2.5.1. We also computed the correlations of the PC scores (PCSs) with 10-year CVD risk. For our main analysis involving the PCSs, we computed BFs for all linear regression models where FTP SUVR was the outcome and any combination of the following covariates constituted the explanatory variables: age, sex, *APOE* ε4 status, score along the first principal component (PCS_1_), PCS_2_, and PCS_3_.

#### 2.5.4 Bayesian power analysis

We performed a Bayesian power analysis using simulations to examine the probability of correctly finding support for the alternative hypothesis (defined as BF ≤ 1/3) and the probability of incorrectly finding support for the null hypothesis (defined as BF ≥ 3) when in fact there was a medium group difference (defined as a Cohen’s *d* equal to 0.5) or a weak correlation (defined as a Pearson’s correlation of 0.3). The former probability can be considered as the Bayesian version of statistical power. The latter is related to the probability of a Type II error in the frequentist framework, which is the probability of falsely not rejecting the null hypothesis. Note that unlike in the frequentist framework where statistical power and the probability of Type II error add up to 1, in this Bayesian approach, the two probabilities we describe based on BFs may add up to less than 1 due to the presence of a range of BFs that are considered inconclusive evidence towards one hypothesis versus the other (namely, the interval 1/3 < BF < 3). To calculate these two probabilities for the t-test analyses, we generated *n*_PiB-_ observations from a normal distribution with zero mean and unit standard deviance and *n*_PiB+_ observations from a normal distribution with mean equal to 0.5 and unit standard deviance, where *n*_PiB-_ and *n*_PiB+_ are the numbers of participants in the PiB- and PiB+ groups, respectively. We then applied the Bayesian t-test with the same prior used in our main analysis and calculated the BF. For the correlation test analyses, we generated *n*_PiB-_ + *n*_PiB+_ observations from a bivariate normal distribution with zero mean and covariance equal to 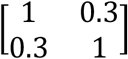. We then applied the Bayesian correlation test with the same prior used in our main analysis and calculated the BF. We obtained 100,000 values for each of these two BFs by repeating these simulations. The probability of correctly finding support for the alternative hypothesis was determined as the fraction of simulations with BF ≤ 1/3, and the probability of incorrectly finding support for the null hypothesis was determined as the fraction of simulations with BF ≥ 3.

#### 2.5.5 Software

We conducted all statistical analyses in R (https://cran.r-project.org, version 4.0.3) (R Core Team, 2020). We used the tidyverse (Wickham et al., 2019) package for data wrangling, factoextra to perform PCA (Kassambara and Mundt, 2020), BayesFactor to compute BFs (Morey and Rouder, 2018), ggthemes (Arnold, 2021), ggpubr (Kassambara, 2020), and wesanderson (Ram and Wickham, 2018) to generate plots, gtsummary (Sjoberg et al., 2021) to create the participant characteristics table, and knitr (Xie, 2020) to generate reports.

#### 2.5.6 Code and data availability

Code for performing statistical analyses and generating figures is provided in an open repository (https://gitlab.com/bilgelm/tau_vascular). Data from the BLSA are available upon request from the BLSA website (https://www.blsa.nih.gov). All requests are reviewed by the BLSA Data Sharing Proposal Review Committee. A deidentified version (with age bands instead of continuous age and excluding *APOE* ε4 status) of the BLSA data used in these analyses is publicly available (https://doi.org/10.7910/DVN/JOMOEN).

## 3 Results

The sample consisted of 87 cognitively normal participants, whose characteristics are summarized in Table 1.

**Table 1.**
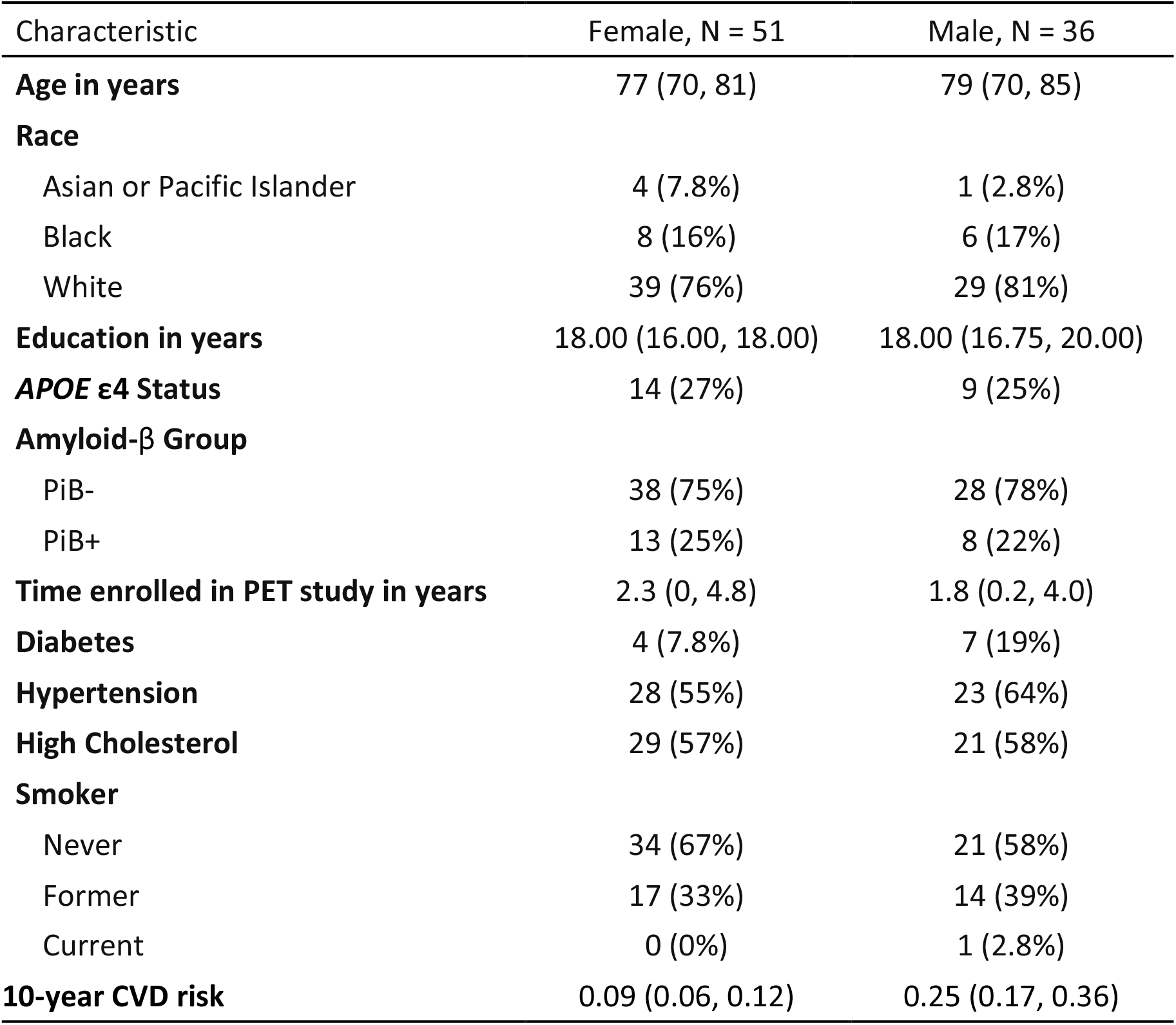
Participant demographics. We report the median (interquartile range) for continuous variables, and N (%) for categorical variables. Cardiovascular disease (CVD) risk is the estimated probability of developing CVD within 10 years calculated using the equations reported by D’Agostino et al. based on the Framingham Heart Study (D’Agostino et al., 2008).

| Characteristic | Female, N = 51 | Male, N = 36 |
| --- | --- | --- |
| **Age in years** | 77 (70, 81) | 79 (70, 85) |
| **Race** |  |  |
| Asian or Pacific Islander | 4 (7.8%) | 1 (2.8%) |
| Black | 8 (16%) | 6 (17%) |
| White | 39 (76%) | 29 (81%) |
| **Education in years** | 18.00 (16.00, 18.00) | 18.00 (16.75, 20.00) |
| **APOE ε4 Status** | 14 (27%) | 9 (25%) |
| **Amyloid-β Group** |  |  |
| PiB- | 38 (75%) | 28 (78%) |
| PiB+ | 13 (25%) | 8 (22%) |
| **Time enrolled in PET study in years** | 2.3 (0, 4.8) | 1.8 (0.2, 4.0) |
| **Diabetes** | 4 (7.8%) | 7 (19%) |
| **Hypertension** | 28 (55%) | 23 (64%) |
| **High Cholesterol** | 29 (57%) | 21 (58%) |
| **Smoker** |  |  |
| Never | 34 (67%) | 21 (58%) |
| Former | 17 (33%) | 14 (39%) |
| Current | 0 (0%) | 1 (2.8%) |
| **10-year CVD risk** | 0.09 (0.06, 0.12) | 0.25 (0.17, 0.36) |

### 3.1 Results based on 10-year CVD risk

#### 3.1.1 Sanity checks

We found a positive correlation between CVD risk and log-transformed total WML ratio (Pearson’s correlation = 0.24, 95% CI = 0.03–0.43, p = 0.03) (Figure 1). PiB+ individuals had higher FTP SUVR in both the EC (difference in means = 0.086, 95% CI = 0.0040–0.17, p = 0.04) and the ITG (difference in means = 0.092, 95% CI = 0.041–0.14, p = 0.0009). There was a positive correlation between mean cortical PiB DVR and FTP SUVR in both the EC (Pearson’s correlation = 0.32, 95% CI = 0.12 – 0.50, p = 0.003) and the ITG (Pearson’s correlation = 0.33, 95% CI = 0.13 – 0.51, p = 0.002).

**Figure 1.**
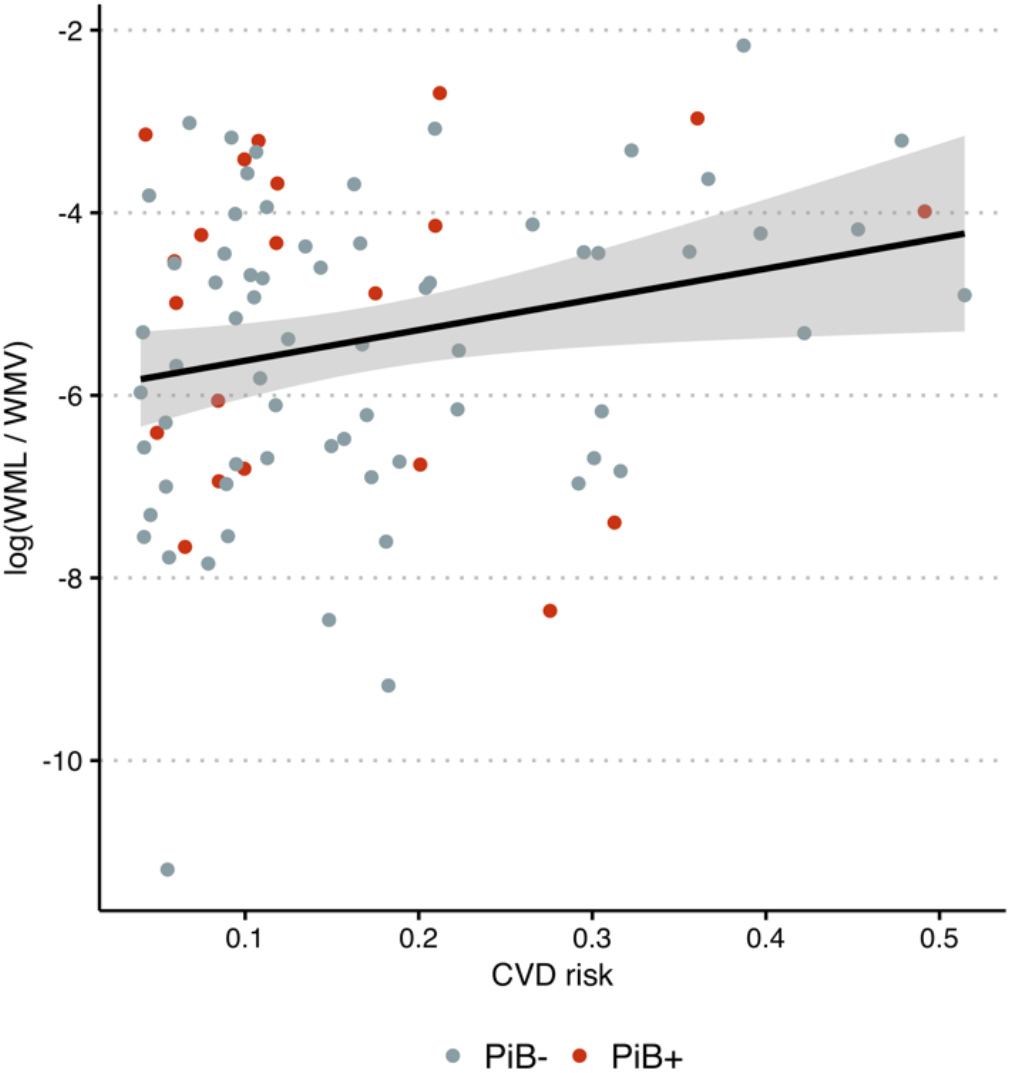
Log-transformed white matter lesion load (WML) to white matter volume (WMV) ratio versus cardiovascular disease (CVD) risk. Color indicates amyloid-β status determined using ^11^C-Pittsburgh compound B (PiB) positron emission tomography.

#### 3.1.2 Main analysis

We did not find a statistically significant correlation between CVD risk and FTP SUVR in either of the regions considered (Pearson’s correlation = -0.11, 95% CI = -0.32 – 0.099, p = 0.3 for EC; Pearson’s correlation = -0.022, 95% CI = -0.23 – 0.19, p = 0.8 for ITG) (Figure 2). The data provided over two- and four-fold evidence towards the null hypothesis that there is no correlation versus the alternative that there is a non-zero correlation between CVD risk and FTP SUVR in the EC (BF = 2.4) and the ITG (BF = 4.0), respectively (Table 2). Similarly, the difference in CVD risk between Aβ groups was small and not statistically significant (difference in means = -0.018, 95% CI = -0.078 – 0.043, p = 0.56) and this finding was supported by a correlation analysis between continuous mean cortical PiB DVR and CVD risk (Pearson’s correlation = -0.038, 95% CI = -0.25 – 0.17, p = 0.72). We did not find a difference in the variance of CVD risk by Aβ group (F_65,20_ = 1.05, p = 0.94). The data provided over three-fold evidence towards the null hypothesis that there is no difference in the mean CVD risk between Aβ groups (BF = 3.4) and the null hypothesis that there is no correlation between mean cortical PiB DVR and CVD risk (BF = 3.8). The sensitivity analysis results for these Bayesian analyses, in which different values of the prior scale parameters are examined, are presented in Figure 3.

**Table 2.** Summary of Bayes factors (BF) for statistical hypotheses regarding the relationship between 10-year cardiovascular disease (CVD) risk as determined by the Framingham Risk Score and amyloid-β and tau pathology as assessed by PiB and FTP PET, respectively. BF is the relative evidence provided by the data towards the null versus the alternative hypothesis.

| Alternative hypothesis | Model | FTP region | BF |
| --- | --- | --- | --- |
| Difference in mean CVD risk by PiB group $\neq 0$ | t-test | – | 3.4 |
| Correlation of CVD risk and PiB DVR $\neq 0$ | Pearson's correlation | – | 3.8 |
| Correlation of CVD risk and FTP SUVR $\neq 0$ | Pearson's correlation | EC | 2.4 |
|  |  | ITG | 4.0 |
| FTP SUVR $\sim 1 + \epsilon 4 + \text{PiB group} + \text{CVD risk}$, where regression coefficient for CVD risk $\neq 0$ | Linear regression | EC | 2.0 |
|  |  | ITG | 3.2 |
| FTP SUVR $\sim 1 + \epsilon 4 + \text{PiB group} + \text{CVD risk} + \text{CVD risk} \times \text{PiB group}$, where regression coefficients for CVD risk and CVD risk × PiB Group interaction $\neq 0$ | Linear regression | EC | 4.7 |
|  |  | ITC | 5.4 |

Abbreviations: CVD cardiovascular disease, BF Bayes factor, DVR = distribution volume ratio, FTP SUVR $^{18}\text{F}$ -flortaucupir standardized uptake value ratio, PiB Pittsburgh compound B, $\epsilon 4$ Apolipoprotein $\epsilon 4$ allele carrier, EC entorhinal cortex, ITG inferior temporal gyrus.

**Figure 2.**
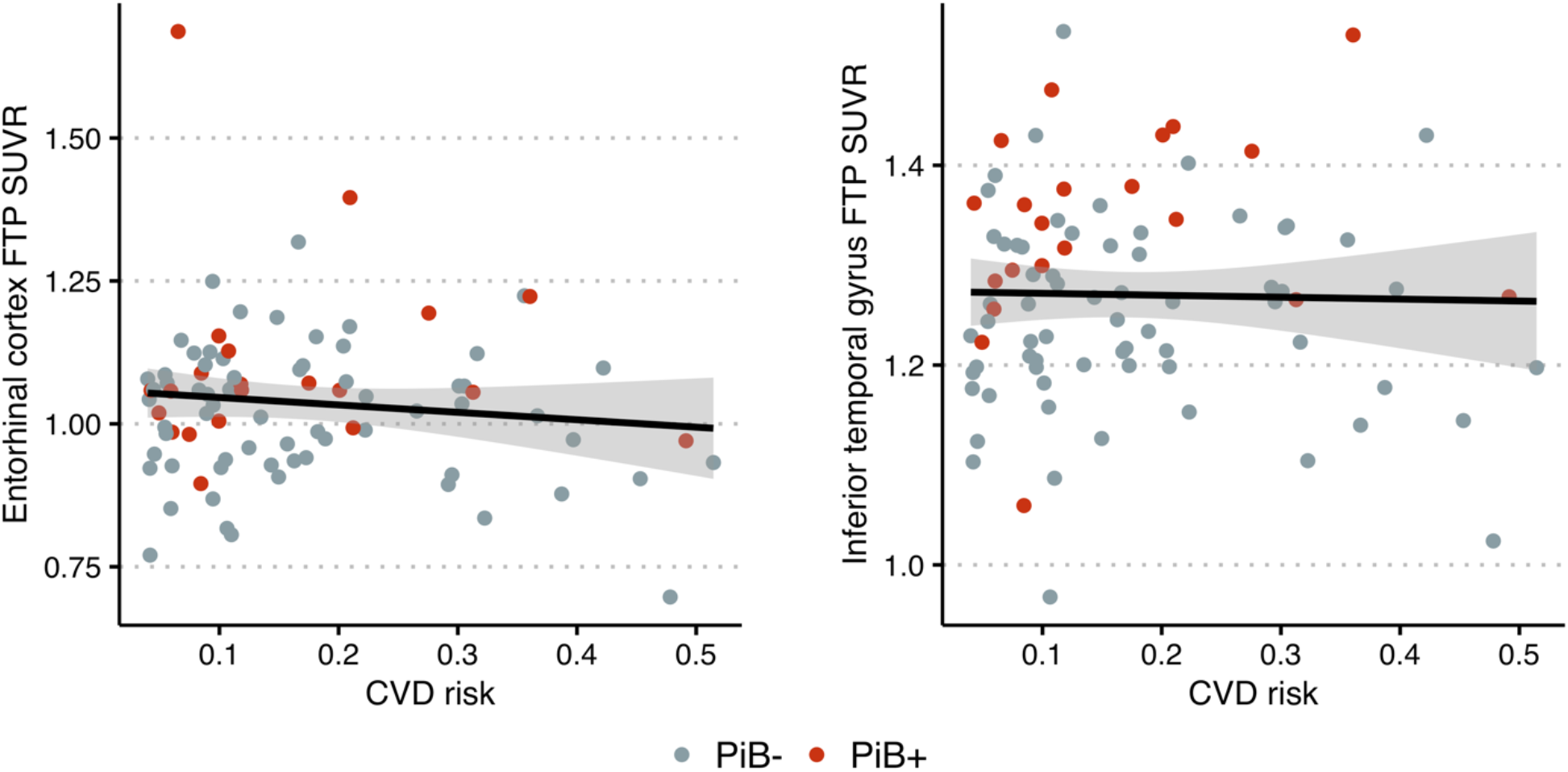
Scatter plots show no relationship in cognitively normal older adults between cardiovascular disease (CVD) risk and ^18^F-flortaucipir (FTP) standardized uptake value ratio (SUVR) in the entorhinal cortex (left) and inferior temporal gyrus (right). Color indicates Aβ status determined using ^11^C-Pittsburgh compound B (PiB) positron emission tomography.

**Figure 3.**
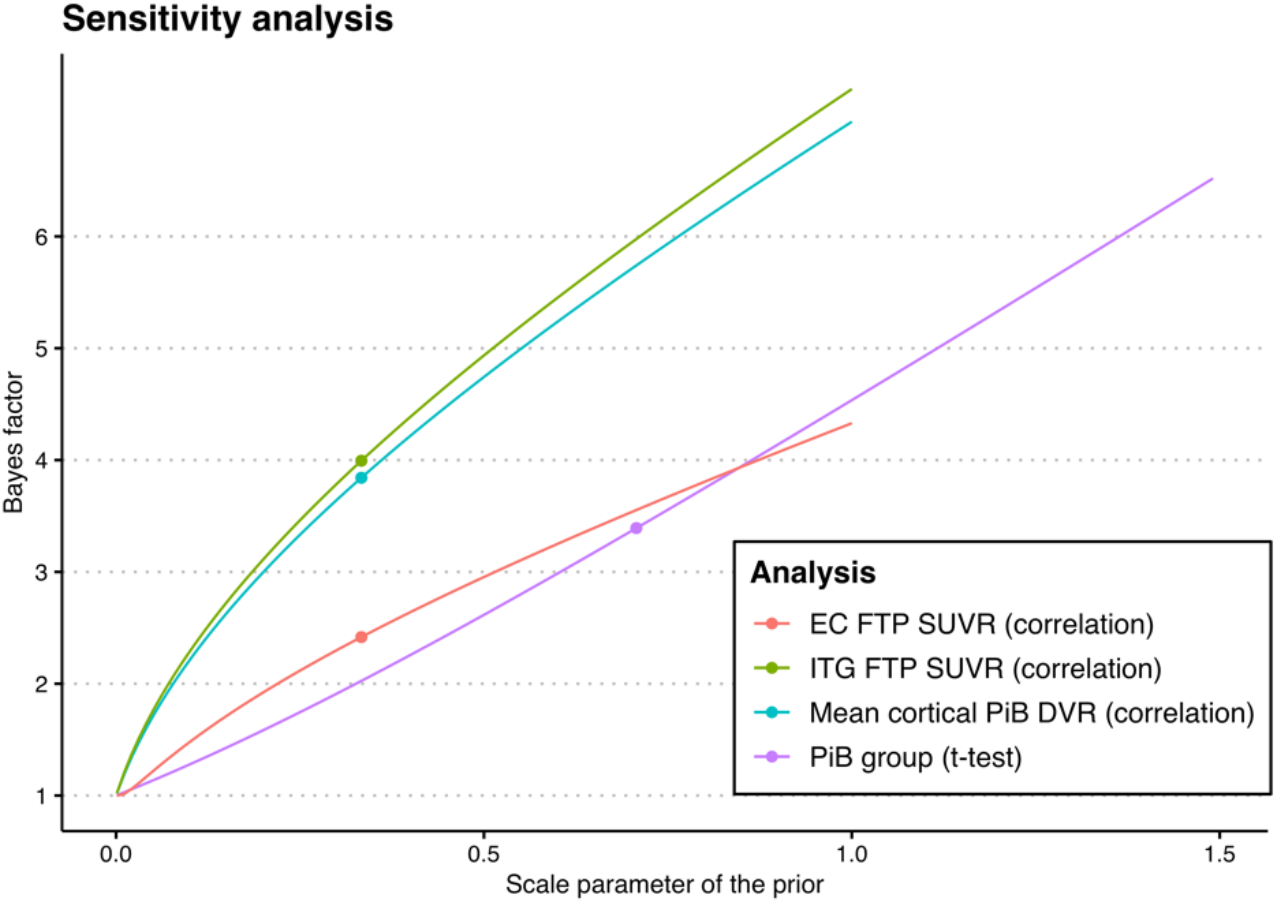
Illustration of the Bayes factors that correspond to different values of the prior scale parameters. Red and green curves correspond to the correlation test between cardiovascular disease risk and ^18^F-flortaucipir (FTP) standardized uptake value ratio (SUVR) in the entorhinal cortex (EC) and inferior temporal gyrus (ITG), respectively, purple curve corresponds to the t-test assessing differences in mean cardiovascular disease risk between Aβ groups, and blue curve corresponds to the correlation test between cardiovascular disease risk and mean cortical ^11^C-Pittsburgh compound (PiB) distribution volume ratio (DVR). Dots indicate the operating points that correspond to medium-width priors.

The data provided more than two-fold evidence towards the linear regression models without the CVD risk term compared to the models with the CVD risk term (BF = 2.0 for EC, 3.2 for ITG). The models that additionally included an interaction between CVD risk and Aβ group were also less favorable compared to the null model that did not include any terms involving CVD risk (BF = 4.7 for EC, 5.4 for ITG).

### 3.2 Results based on principal component scores

#### 3.2.1 Sanity checks

The correlation coefficients between log-transformed total WML ratio and PCSs were not statistically significant (all p > 0.2) (Supplementary Figure 3). On the other hand, there was a statistically significant weak correlation between 10-year CVD risk and each of the PCSs (−0.34 for PCS_1_, 0.32 for PCS_2_, and -0.24 for PCS_3_; all p < 0.03) (Supplementary Figure 4).

#### 3.2.2 Main analysis

We did not observe a statistically significant correlation between FTP SUVR in the EC or ITG and any of the three PCSs (Supplementary Figure 5), nor did we find a difference between Aβ groups in any of the PCSs or a correlation between mean cortical PiB DVR and any of the PCs. For EC FTP SUVR, the model with the highest BF included only Aβ status as an explanatory variable, and this model constituted our null hypothesis that none of the PCSs are associated with EC FTP SUVR. The model with the second highest BF additionally included PCS_3_ (i.e., scores along the third principal component), and this model constituted our alternative hypothesis. The data provided more than two-fold evidence towards the former model (BF = 2.2). For ITG FTP SUVR, the model with the highest BF included sex and Aβ status as explanatory variables. The model with the second highest BF additionally included PCS_3_, but the data provided almost three-fold evidence towards the former model (BF = 2.9).

### 3.3 Bayesian power analysis results

The Bayesian power analysis showed that the probability of correctly finding support for the alternative hypothesis (comparable to statistical power in the frequentist framework) was 44% for the t-test and 70% for the correlation test. The probability of incorrectly finding support for the null hypothesis was 9% for the t-test and 2% for the correlation test. Distributions of the BFs across simulations are shown in Supplementary Figure 6.

## 4 Discussion

In this cross-sectional study of cognitively normal individuals, vascular risk, quantified using the Framingham 10-year CVD risk or via PCA, was not associated with Aβ or tau pathology. We consistently found over two-fold evidence in support of a lack of an association versus a non-zero association between vascular risk and neuropathology. In addition, we did not find a moderation effect of Aβ on the relationship between vascular risk and tau pathology. Although disagreeing with our original hypothesis that higher vascular risk would be associated with greater levels of Aβ and tau, our findings suggest that vascular risk and AD neuropathology may constitute separate pathways and independently contribute to the development of cognitive impairment.

Our findings agree with studies among cognitively normal individuals that did not find a correlation between vascular risk and Aβ (Bos et al., 2019; Glodzik et al., 2014; Laws et al., 2017; Marchant et al., 2012; Pettigrew et al., 2020; Rabin et al., 2018; Vassilaki et al., 2016; Vemuri et al., 2015; Walters et al., 2018) or tau (Köbe et al., 2020; Pettigrew et al., 2020). On the other hand, our findings differ from those reported by Rabin et al. suggesting that vascular risk and Aβ pathology work synergistically to accelerate the manifestation of tau pathology among cognitively normal individuals (Rabin et al., 2019). The discrepancy might be due to differences in sample composition: although both studies assessed vascular risk with the Framingham 10-year CVD risk score, Rabin et al. found a wider range (4% - 74%) of CVD risk in their sample compared to ours (4% - 51%). Additionally, their sample consists of a higher percentage of APOE e4 carriers (31% vs. 26%), who on average have been shown to have greater vascular risk (Mielke et al., 2011) and AD neuropathology (Baek et al., 2020).

The role of vascular risk and neuropathology in AD has been widely studied. A review by (Zlokovic, 2011) presents several studies supporting a two-hit hypothesis, which suggests that vascular risk leads to the development of dementia through the combination of cerebrovascular disease, the reduction of Aβ clearance and increased production of Aβ. The increase of Aβ accelerates the neurodegenerative pathway to dementia, including the formation of neurofibrillary tau tangles. While our results do not refute this hypothesis, they provide evidence against such an involvement of vascular risk in the preclinical stage of AD. The absence of an association between vascular risk and tau pathology in our study supports the possibility that vascular risk impacts the development of clinical AD independent of neuropathology. Indeed, studies have found that vascular risk and Aβ pathology have additive, not synergistic, effects on cognitive decline (Pettigrew et al., 2020; Vemuri et al., 2015). Several other studies have also found that vascular risk acts independent of AD biomarkers to contribute to cognitive decline (Marchant et al., 2012; Rosano et al., 2007). It is possible that the effect of vascular risk on the development of AD is due to vascular risk factors leading to concomitant cerebrovascular disease and vascular brain injury later in life, which lead to cognitive decline and dementia (Chui et al., 2012).

Our study has several limitations. First, BLSA participants are generally well-educated and primarily White, which impacts the generalizability of our results to the larger population. Second, participants who have severe cardiovascular and cerebrovascular disease are ineligible for enrollment into the PET substudy. These enrollment exclusion criteria may have resulted in a sample consisting of individuals with better overall vascular health, thus limiting the applicability of our results to those with higher vascular risk. However, participants are not excluded from the study if they develop these conditions after enrollment. The median time enrolled in the PET substudy prior to FTP PET for our sample was about 2 years. Therefore, some participants may have developed cardiovascular and cerebrovascular conditions, such as stroke or infarcts, after enrollment. Third, our study was based on cross-sectional data. Longitudinal studies will be needed to better understand the relationship between vascular risk and tau accumulation over time. Fourth, we did not examine individual vascular risk factors in relation to Aβ or tau pathology. This is an important question that would be better addressed in larger samples to allow for multiple comparison correction. Finally, our sample size was small (n=87), which did not yield at least 80% statistical power for detecting weak correlations (i.e., with magnitude below 0.3). However, the probability of finding at least moderate evidence toward the null hypothesis (i.e., BF ≥ 3) under the assumption of a true correlation with magnitude 0.3 was only 2%, which suggests that if there were indeed at least a weak correlation between CVD risk and Aβ or tau pathology among cognitively normal older adults, it would have been quite unlikely to find the level of support towards the null hypothesis that we reported.

A notable strength of this study is the use of a combination of objective, subjective, dichotomous, and continuous measures of vascular risk. For instance, in addition to using self-reported diagnosis of vascular health conditions, hypertension, diabetes, and high cholesterol, we also considered medication use and suprathreshold measurements of continuous variables such as SBP, DPB, fasting glucose, and total cholesterol in determining diagnosis. This allowed us to obtain robust measures of vascular risk within our sample. Additionally, the calculation of Bayes factors allowed us to directly assess the degree of evidence provided by the data in support of the lack of an association between vascular risk and neuropathology, which is not possible under a null hypothesis significance testing framework, where one can only reject or fail to reject the null hypothesis.

## 5 Conclusions

While there is evidence supporting the concept that vascular risk may promote and interact synergistically with Aβ and tau pathology, our findings are more in accord with an independent relationship, at least among cognitively normal individuals. Nevertheless, as cerebrovascular and cardiovascular disease play a significant role in the development of dementia, focusing on the vascular risk factors which lead to these diseases may be a favorable approach for designing interventions and treatments aimed at reducing risk of dementia. Future studies with larger samples and longitudinal follow-up are needed to better elucidate the relationship between vascular risk and neuropathology and their interplay in relation to cognitive outcomes.

## Data Availability

Data from the BLSA are available upon request from the BLSA website (https://www.blsa.nih.gov). All requests are reviewed by the BLSA Data Sharing Proposal Review Committee. A deidentified version (with age bands instead of continuous age and excluding APOE ε4 status) of the BLSA data used in these analyses is publicly available (https://doi.org/10.7910/DVN/JOMOEN).

https://doi.org/10.7910/DVN/JOMOEN

## Abbreviations

Aβ: Amyloid-beta
AD: Alzheimer’s disease
*APOE*: Apolipoprotein E (gene)
BF: Bayes factor
BLSA: Baltimore Longitudinal Study of Aging
CI: confidence interval
CSF: cerebrospinal fluid
CVD: Cardiovascular disease
DVR: Distribution volume ratio
EC: entorhinal cortex
FLAIR: Fluid attenuated inversion recovery
FTP: ^18^F-flortaucipir
FWHM: Full-width at half-maximum
ITG: inferior temporal gyrus
HRRT: High resolution research tomograph
MPRAGE: Magnetization-prepared rapid gradient echo
MRI: Magnetic resonance imaging
PC: Principal component
PCA: Principal component analysis
PCS: Principal component score
PET: Positron emission tomography
PiB: ^11^C-Pittsburgh compound B
SBP: systolic blood pressure
DBP: diastolic blood pressure
SUVR: Standardized uptake value ratio
TE: echo time
TR: repetition time
WML: white matter lesion load
WMV: white matter volume

## Acknowledgments

This study was supported by the Intramural Research Program of the National Institute on Aging, NIH. We thank the Johns Hopkins PET facility staff, the NIA 3T MRI facility staff, Wendy Elkins for her assistance with coordination and data collection for the BLSA PET studies, the Center for Biomedical Image Computing and Analytics for providing MUSE labels and their contributions to MRI analysis, and Avid Radiopharmaceuticals for enabling the use of the ^18^F-flortaucipir tracer and providing the precursor. Avid Radiopharmaceuticals did not provide direct funding for any of the PET studies nor personnel and were not involved in data analysis or interpretation.

## Supplementary Material

Principal component analysis (PCA)

We converted each of the eight vascular risk-related measurements (BMI, MAP, HDL, LDL, total cholesterol, triglycerides, fasting glucose, smoking status [current smoker vs. not]) in the larger BLSA data set (n=1588 participants) to *z*-scores. We then split the data into training (n=794 participants) and testing (n=794 participants) sets. We verified that the training and testing sets had comparable distributions for age, sex, and the vascular variables by comparing continuous variables using t-tests and categorical variables using Fisher’s exact test. We did not find any statistically significant differences between training and testing sets. We performed PCA separately on each set to compute principal components in the training set (PC_train_) and in the testing set (PC_test_). We oriented the PCs (and the corresponding scores) such that LDL had a positive loading on each component. We kept the first three components in each data set. The first three components explained 70.3% and 72.1% of the variance in the training and testing sets, respectively. The loadings for each principal component were similar between the training and testing sets (Supplementary Table 1). To quantify the level of similarity, we calculated scores for each participant in the testing set by projecting their measurements onto PC_train_, and assessed the Pearson’s correlation between the scores based on PC_train_ and scores based on PC_test_ for each participant in the testing set. We found high correlations along each principal component (Pearson’s correlation = 0.90 along 1^st^component, 0.89 along 2^nd^component, and 0.98 along 3^rd^component; all p < 0.001) (Supplementary Figure 1). Based on these results, we concluded that PCA of these 8 vascular risk-related measurements yields sufficiently replicable results. The principal components computed in the training set were used to calculate scores for each of the 87 participants included in this study. The difference in the distribution of the scores among these 87 participants and the distribution observed in the training set was not statistically significant for any PC (Kolmogorov-Smirnov test all p > 0.3), suggesting that participants in the BLSA tau PET substudy have vascular risk profiles that are comparable to those observed in the larger BLSA sample (Supplementary Figure 2).

---

**Supplementary Table 1.** Principal component (PC) loadings in the training and testing sets.

|  | Training |  |  | Testing |  |  |
| --- | --- | --- | --- | --- | --- | --- |
|  | PC 1 | PC 2 | PC 3 | PC 1 | PC 2 | PC 3 |
| BMI | -0.168 | 0.468 | -0.145 | -0.337 | 0.31 | -0.216 |
| MAP | -0.103 | 0.147 | -0.925 | -0.132 | 0.151 | -0.865 |
| HDL | -0.329 | 0.383 | 0.284 | -0.516 | 0.238 | 0.3 |
| LDL | 0.582 | 0.328 | 0.028 | 0.373 | 0.565 | 0.042 |
| Cholesterol | 0.657 | 0.242 | -0.06 | 0.492 | 0.5 | -0.022 |
| Triglycerides | -0.025 | 0.529 | 0.195 | -0.295 | 0.464 | 0.305 |
| Glucose | -0.286 | 0.409 | -0.007 | -0.367 | 0.198 | -0.144 |
| Smoking | -0.004 | 0.005 | 0.003 | -0.005 | 0.001 | 0.001 |
| *Variance explained* | 28.5% | 27.9% | 14% | 29.7% | 27.5% | 15% |

**Supplementary Figure 1.**
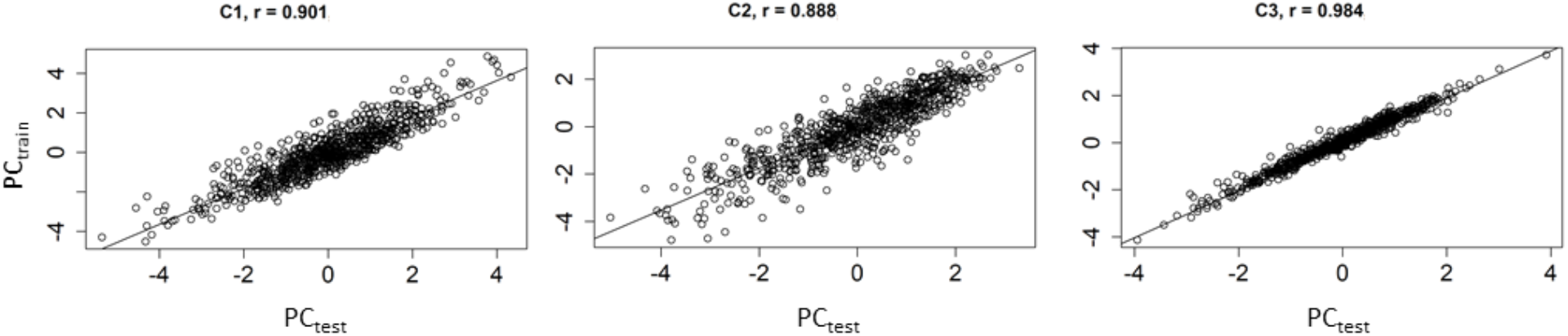
Comparison of the component scores generated across the two data sets. Correlations are between scores based on the projection of test data onto PC_train_ and scores from PCA_test_.

**Supplementary Figure 2.**
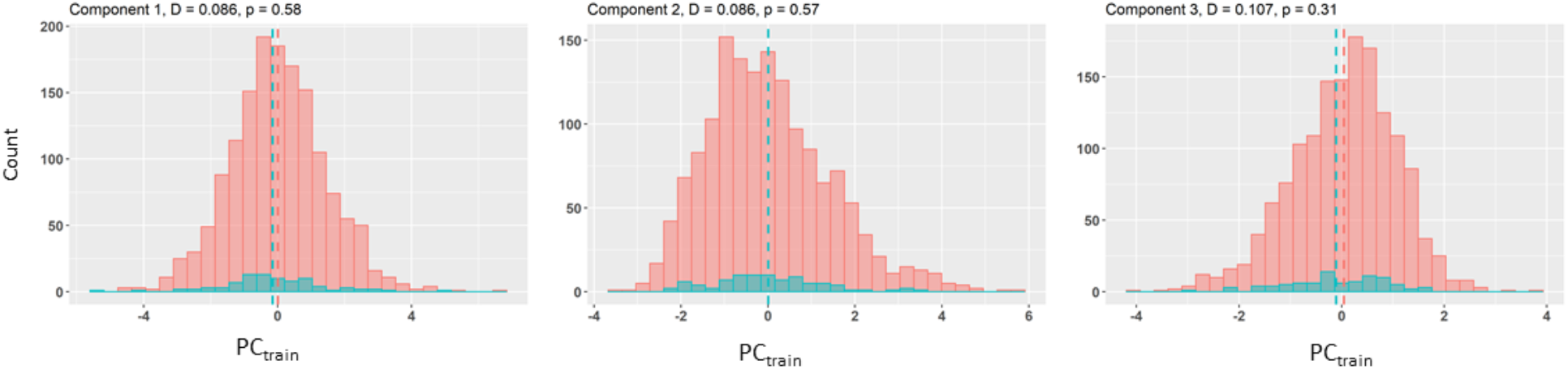
Histograms showing the scores for the 1501 non-FTP PET BLSA participants (pink) and the 87 FTP PET participants (blue). Scores are based on PC_train_. The vertical dashed lines represent the group means. The Kolmogorov-Smirnov test statistic (D) and its corresponding p value are indicated above each panel.

**Supplementary Figure 3.**
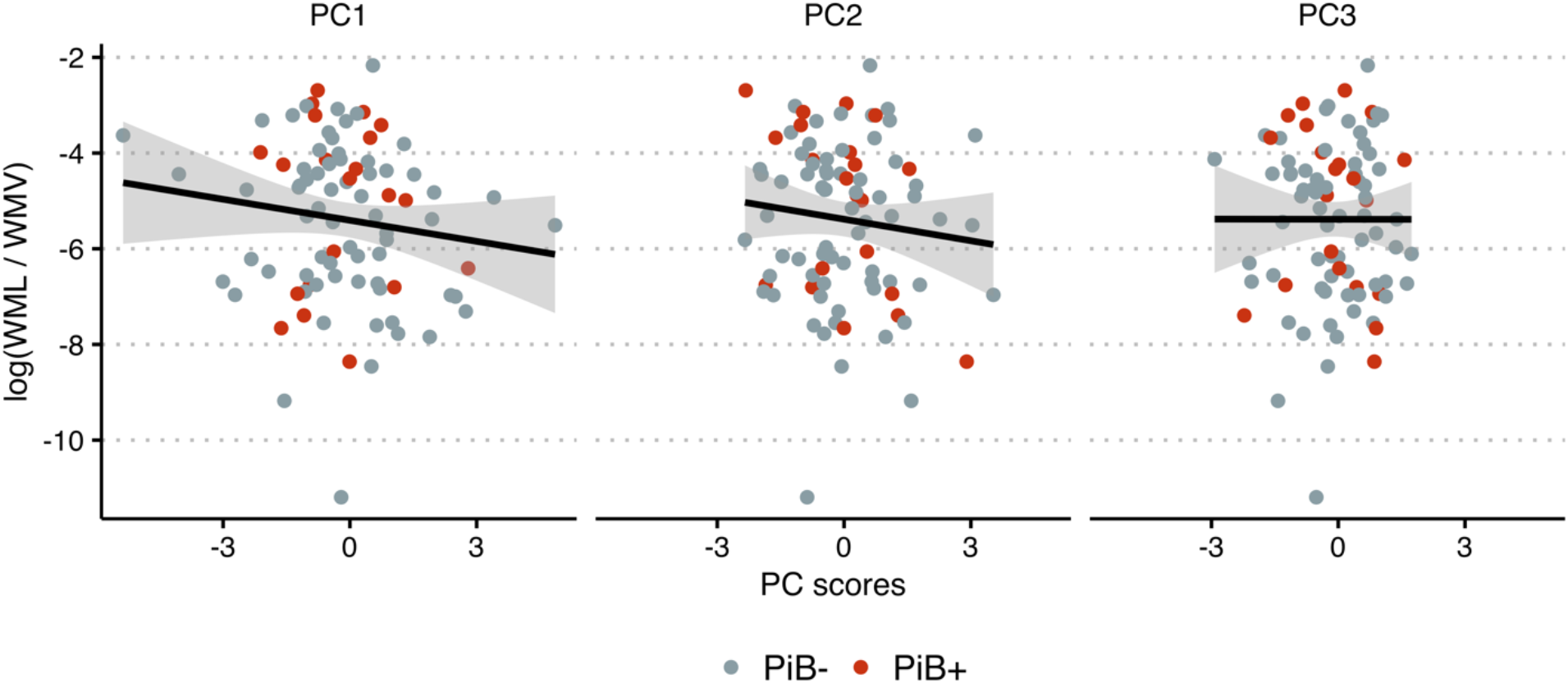
Log-transformed white matter lesion load (WML) to white matter volume (WMV) ratio versus principal component (PC) scores. Color indicates amyloid-β status determined using ^11^C-Pittsburgh compound B (PiB) positron emission tomography.

**Supplementary Figure 4.**
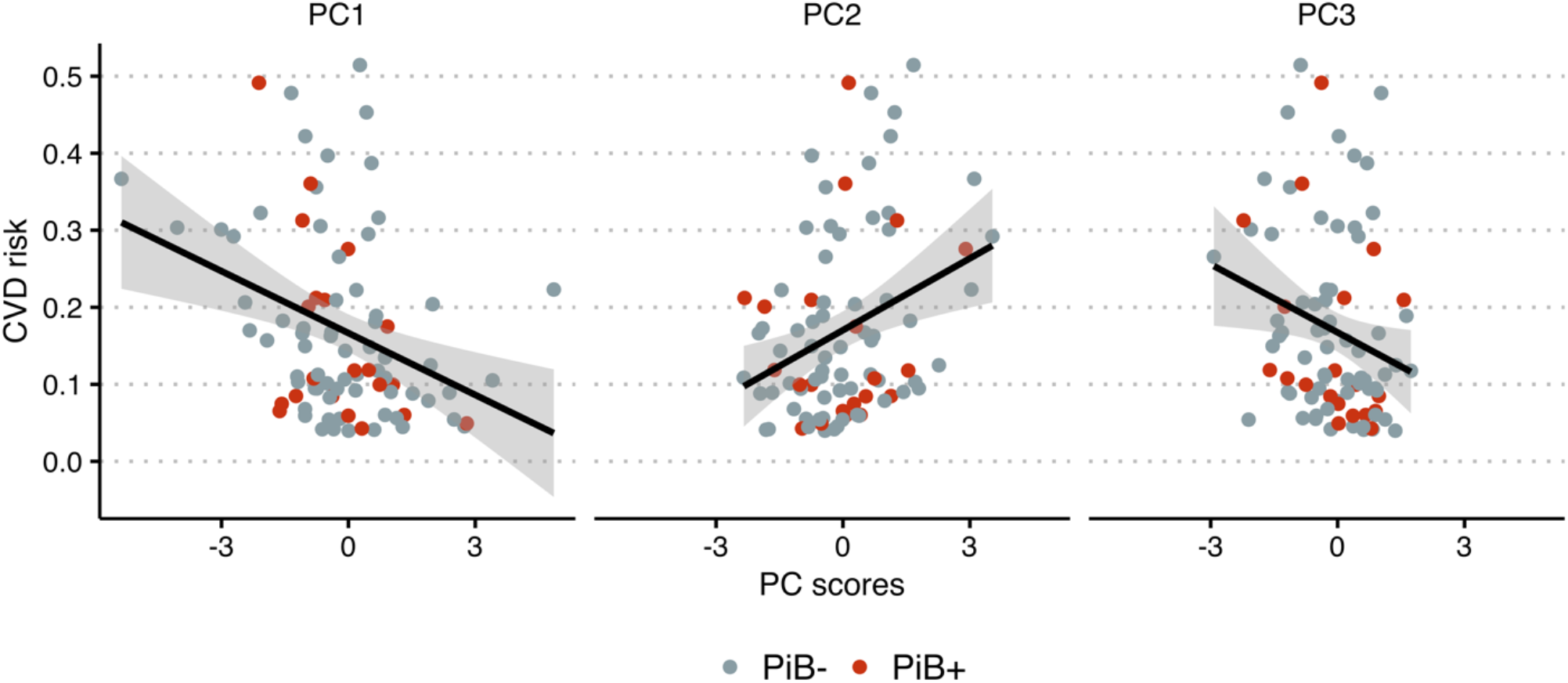
10-year cardiovascular disease (CVD) risk versus principal component (PC) scores. Color indicates amyloid-β status determined using ^11^C-Pittsburgh compound B (PiB) positron emission tomography.

**Supplementary Figure 5.**
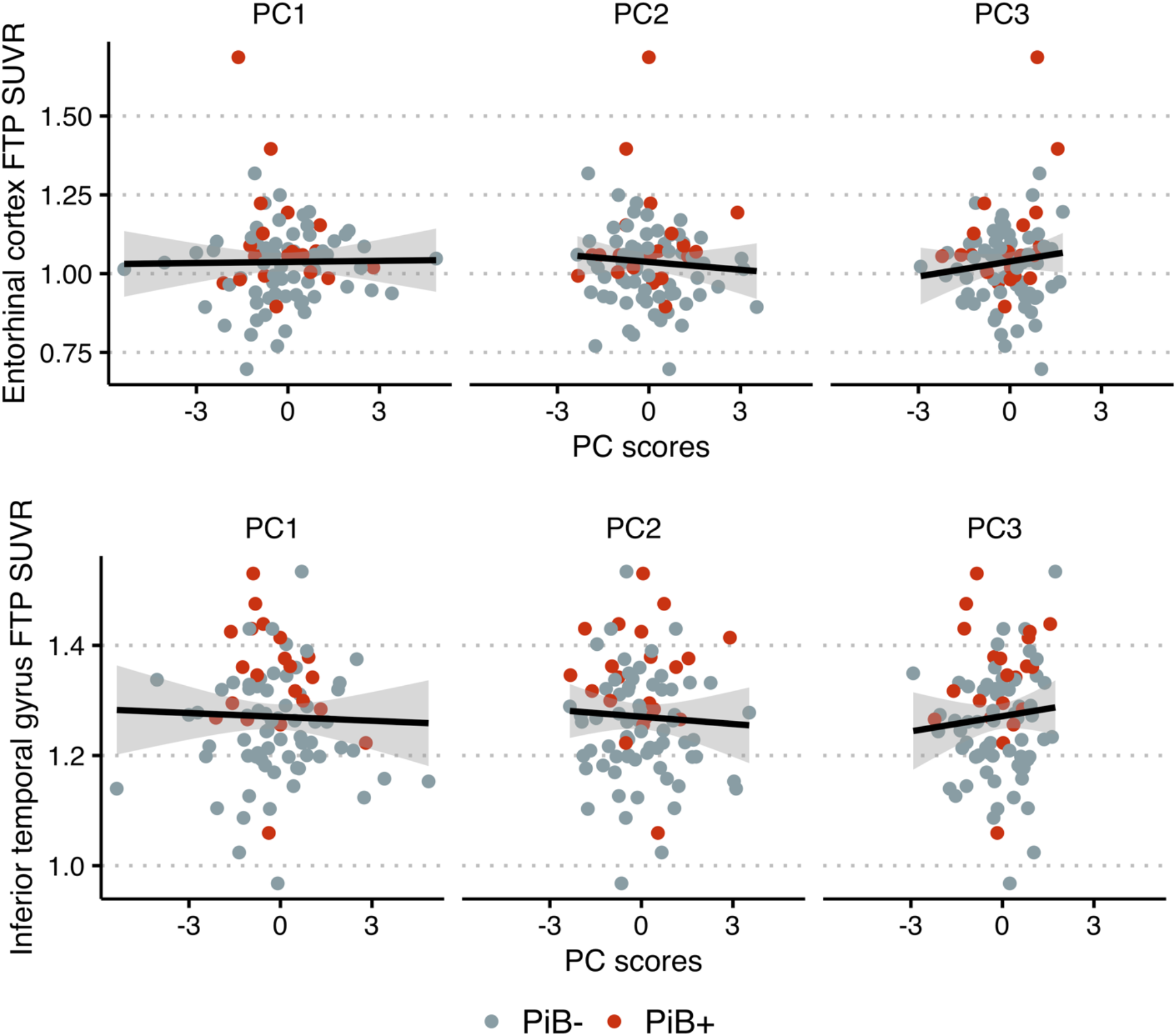
Scatter plots show no relationship in cognitively normal older adults between principal component (PC) scores and FTP SUVR in the entorhinal cortex (left) and inferior temporal gyrus (right). Color indicates amyloid-β status determined using ^11^C-Pittsburgh compound B (PiB) positron emission tomography.

**Supplementary Figure 6.**
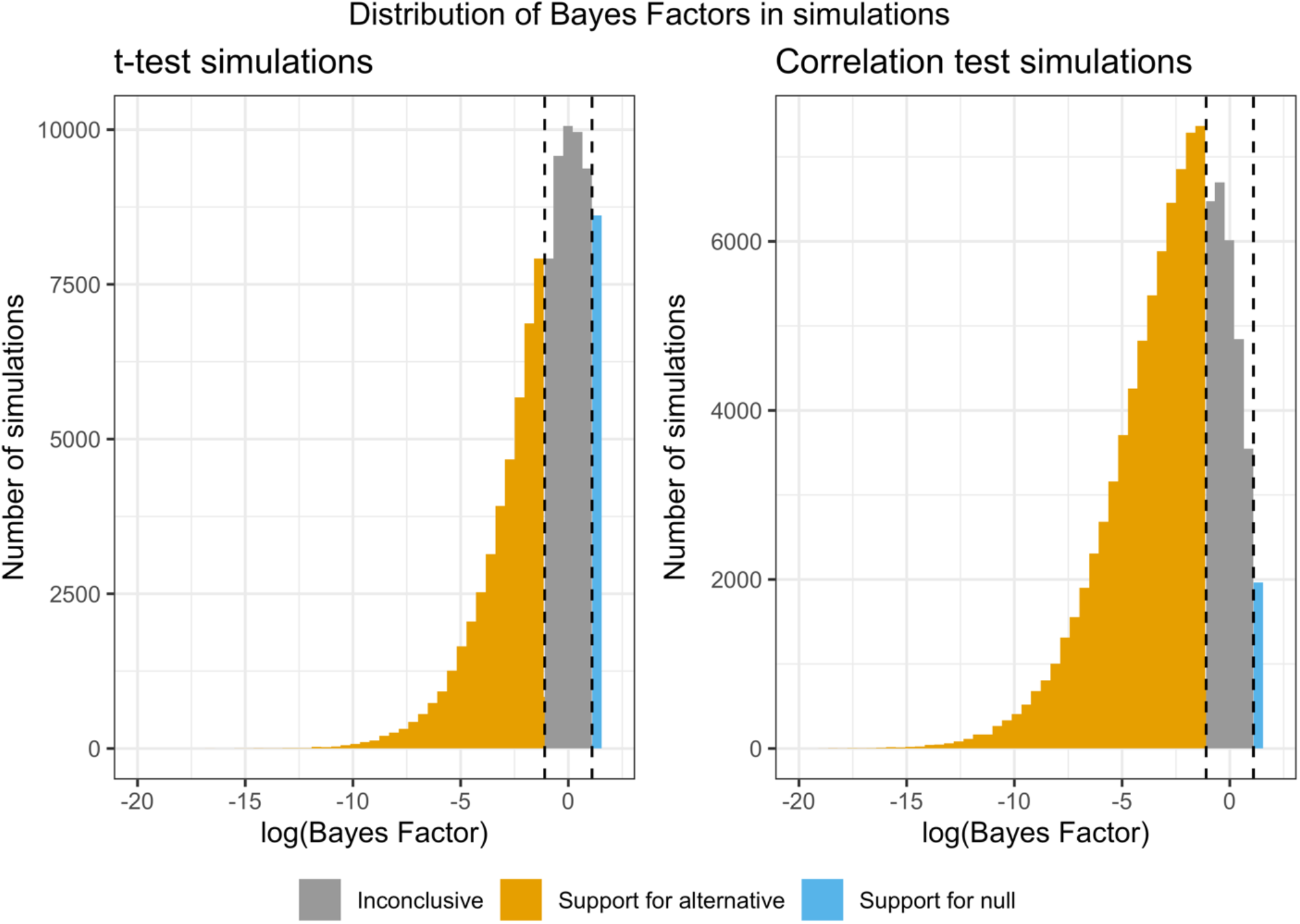
Distribution of Bayes factors calculated in simulations where data were generated according to the alternative hypothesis. In t-test simulations, the alternative hypothesis was that the effect size (Cohen’s *d*) was 0.5. In correlation test simulations, the alternative hypothesis was that the correlation was 0.3. Since data were generated according to the alternative hypothesis, concluding that the data provide evidence towards the null (i.e., effect size or correlation equals zero) rather than the alternative hypothesis would be erroneous. These simulations demonstrate that the probability of such erroneous conclusions (as indicated in blue) is quite low for both the t-tests and the correlation tests: 9% and 2%, respectively.

## Footnotes

* In the Bayesian t-test, variance was assumed to be equal between amyloid groups. This assumption was assessed using an *F*-test comparing group variances

## Notes

### Competing Interest Statement

The authors have declared no competing interest.

### Summary of Updates

Added analyses using continuous amyloid level instead of dichotomous amyloid status; added Bayesian power analysis; updated data availability.

